# Building and validating 5-feature models to predict preeclampsia onset time from electronic health record data

**DOI:** 10.1101/2023.03.23.23287655

**Authors:** Hailey K Ballard, Xiaotong Yang, Aditya Mahadevan, Dominick J Lemas, Lana X Garmire

## Abstract

**Background:** Preeclampsia is a potentially fatal complication during pregnancy, characterized by high blood pressure and presence of proteins in the urine. Due to its complexity, prediction of preeclampsia onset is often difficult and inaccurate.

**Methods:** This study aims to create quantitative models to predict the onset gestational age of preeclampsia using electronic health records. We retrospectively collected 1178 preeclamptic pregnancy records from the University of Michigan Health System(UM) as the discovery cohort, and 881 records from the University of Florida Health System(UF) as the validation cohort. We constructed two Cox-proportional hazards models with Lasso regularization: one baseline model utilizing maternal and pregnancy characteristics, and the other full model with additional lab results, vital signs, and medications in the first 20 weeks of pregnancy. We built the models using 80% of the UM data and subsequently tested them on the remaining 20% UM data and validated with UF data. We further stratified the patients into high and low risk groups for preeclampsia onset risk assessment.

**Findings:** The baseline model reached C-indices of 0·64 and 0·61 in the 20% UM testing data and the UF validation data, respectively, while the full model increased these C-indices to 0·69 and 0·61 respectively. Both the baseline and full models contain five selective features, among which number of fetuses in the pregnancy, hypertension and parity are shared between the two models with similar hazard ratios. In the baseline model, history of complicated type II diabetes and a mood/anxiety disorder during the first 20 weeks of pregnancy were important. In the full model, maximum diastolic blood pressure in early pregnancy was the predominant feature.

**Interpretation:** Electronic health record data provide useful information to predict gestational age of preeclampsia onset. Stratification of the cohorts using five-predictor Cox-PH models provide clinicians with convenient tools to assess the patients’ onset time of preeclampsia.

**Funding:** This study was supported by grants through the NIEHS, NICHD, NIDDK, and NCATS.

## Introduction

Preeclampsia (PE) is a pregnancy-associated condition characterized by new-onset hypertension and proteinuria, typically diagnosed after 20 weeks of gestation in approximately 3-5% of all pregnancies.^1^ As one of the leading causes of maternal mortality and morbidity worldwide, it is primarily caused by improper placentation and can lead to a more serious condition called eclampsia if left untreated.^2^ Timely identification of PE is a key factor in pregnancy risk management and subsequent treatment. The National Institute for Health and Care Excellence(NICE) recommends identifying and preventatively treating women at high risk for PE (those with preexisting hypertension, chronic kidney disease, insulin-dependent diabetes, and previously early-onset PE) before 13 weeks of gestation.^3^ However, diagnosis of PE is mostly dependent on clinical markers such as blood pressure, urinary protein excretion, mean arterial pressure, and placental growth factor levels, and these markers do not typically show abnormality until after 20 weeks of gestation.

Previous studies have identified some risk factors of PE, including PE in a previous pregnancy, a multifetal pregnancy, chronic hypertension before pregnancy, diabetes before pregnancy, kidney disease, autoimmune disorders, as well as demographic factors including obesity, advanced maternal age, and race.^4^ However, the quantitative importance of these risk factors relative to each other has not been adequately investigated. Additionally, although elevated blood pressure and high levels of protein in the urine are two classic clinical symptoms utilized to diagnose PE, many women develop PE without experiencing at least one of them.^5^ The cumulative effects of some understudied risk factors are unknown. There is an unmet need to provide clinicians with tools to accurately identify when mothers at risk will develop PE or EOPE, which would allow the prescription of preventative treatment before symptoms manifest further. However, many of these risk factors fail to capture the influence on PE onset time - rather, they are considered dichotomous risk factors that simply increase the risk. Haile et al. discuss how maternal age, weight, and history of PE significantly drive PE onset time,^6^ but many additional factors remain undefined.

Computational modeling using large-scale patient-level health data provides opportunities to systematically address all the issues above. However, PE prediction has thus far only been modeled as a classification problem, ignoring the wide range of variations in onset time. A most recent study stratified PE patients by gestational weeks to build classification models, resulting in many models that are difficult for clinicians to choose.^7^ Moreover, these classification models can not predict the onset time for an individual patient, thus failing to assist clinicians in making early decisions on preventative therapy. Unlike all previous attempts to model PE risks, the work here utilizes comprehensive EMR data (demographics, medical history, lab results, vitals, medications, etc.) to explicitly predict the onset gestational age of PE, framed as survival prediction using the Cox-PH model. This approach enables the investigation of risk factors (features) that may affect PE onset time. Features are selected through lasso feature selection and their hazard ratios (HRs) reported. This model outputs risk factors that influence PE development and predicts time to PE diagnosis for patients. Risk factors with a hazard ratio greater than one can help clinicians more accurately identify patients at higher risk for earlier PE development, and a shorter time to PE diagnosis. Additionally, the patients can also be stratified into low-risk and high-risk PE groups, accompanied by differences in risk factors (features). These models will allow clinicians to practically identify when an at-risk mother might develop PE and reveal any features associated with the onset time of PE that are not included in the current guideline.

## Methods

### Data Source

The discovery cohort for this project was obtained from the University of Michigan (UM) Medicine Healthcare System. It is an academic health system that serves Ann Arbor, Michigan and the surrounding areas. The Institutional Review Board of the University of Michigan Medical School (HUM#00168171) approved this usage.The data were downloaded from the Precision Health Analytic platform, a web-based interface to access de-identified EMR data.^8^ All pregnant records (between years 2015 and year 2021) with at least one PE diagnosis, based on the International Classification of Diseases (ICD)-10 codes, were extracted **(Supplementary Table 1**). Patients who were diagnosed with HELLP syndrome, chronic hypertension with superimposed PE, and postpartum PE, were removed from the cohort. We also removed patients transferred from other institutions by deleting patients who did not have an encounter in the University of Michigan system within 20 weeks of the start of their pregnancy. Due to the fact that PE is clinically defined after 20 weeks, all patients with a PE diagnosis before 20 weeks gestation were dropped from the discovery cohort. 1178 pregnancies remained in the UM discovery cohort following this data selection.

Following the same inclusion and exclusion criteria, the validation cohort was generated from the University of Florida Health System (UFHealth) and contained 881 preeclamptic pregnancies from 2015 to 2021. The Integrated Data Repository, as approved by the UF IRB (#201601899) managed the de-identification and transfer of patient data to the researchers.

### EMR Feature Extraction

The EMR provided features, including medical history, diagnoses made during each unique pregnancy, demographics, medications, lab results, and vital signs. The baseline model initially utilized features including age at the start of pregnancy, race, date of pregnancy start, date of the first PE diagnosis, gravidity, parity, and previous history of PE at the trimester it was diagnosed. Additionally, medical histories based on ICD diagnosis codes were extracted using the Elixhauser Comorbidities definitions,^9^ including uncomplicated hypertension, uncomplicated and complicated diabetes (both type I and type II), autoimmune disorders such as systemic lupus erythematosus, hypertension, polycystic ovarian syndrome, mood disorders such as anxiety and depression, and kidney disease. ICD diagnosis codes were also extracted for any diagnosis made within the first 20 weeks of the pregnancy. These codes included the same features as in the medical history, as well as headaches, alcohol or drug use, any infection, smoking, sleep apnea, pregnancy via in vitro fertilization, and a body mass index (BMI) over 35.

For the full model, lab results, vital signs, and medications ordered before 20 weeks of gestation were also added. Based on a literature review of common medications that are prescribed during pregnancy and may be related to PE development, medications captured included benzodiazepine, antacids, nonsteroidal anti-inflammatory drugs (NSAIDs), nasal sprays, acetaminophen, calcium channel blockers, and triptans.^10^ Lab tests captured were those common to a blood panel, including hematocrit, hemoglobin, mean corpuscular hemoglobin, mean corpuscular hemoglobin concentration (MCHC), mean platelet count, mean platelet volume (MPV), white blood cell count (WBCC), red blood cell count, and red cell distribution width. Vital signs included diastolic and systolic blood pressure. In the full model, the time window for labs and vitals was from the start of pregnancy (0 days gestation) to 20 weeks (140 days) gestation. The mean, maximum, minimum, and standard deviation for each lab value were calculated. Patients who didn’t have any encounter within the first twenty weeks (∼15%) were assigned ‘missing’ and the lab and vital values were imputed. All categorical features were converted into binary variables. All numeric variables were log-transformed to adjust for skewness. Each feature in the medical history, clinical diagnosis, and medication categories were computed as a binary category: 1 for presence of a diagnosis, 0 for absence. This method was used to reduce feature dimensionality and improve interpretability.

All analysis was conducted using R 4.2.2^11^. Data cleaning was done using the packages “dplyr”^12^ and “gtsummary”^13^. Model building and cross-validation was conducted using the “glmnet”^14^ package in R. Missing data was imputed using the predictive mean matching algorithm from the R package “mice”.^15^

### Model Construction, Validation and Evaluation

We randomly divided the UM discovery dataset into a training set (80%) and a hold-out testing set (20%). We then constructed a Cox-PH model with LASSO regularization (feature selection) through five-fold cross-validation. The output of the Cox-PH model is the log hazard ratios, also called the prognosis index (PI), which depicts the relative risk of a patient when compared against the baseline hazard of the population. The model is then tested on the hold-out testing set and external UF validation cohort. We constructed the full models using the training cohort in the same way as the baseline model. We evaluated the performance of each model using metrics including the concordance index (C-index) and log-rank p-values. The C-index is a metric to compare the discriminative power of a risk prediction model which describes the frequency of concordant pairs among all pairs of patients included in the model construction.^16^ We used the C-index calculated from the “glmnet”^14^ package. We stratified low-risk and high-risk pregnancies based on the median PI score of the model and plotted the Kaplan-Meier (KM) curves for each risk group. We tested their difference with log-rank test using the training dataset, hold-out testing dataset, and the validation dataset separately in order to evaluate the discriminative power of the model. The log-rank test is a significance test in survival analysis, with the null hypothesis that two groups have identical distributions of survival time. Any log-rank p-value below 0·05 is considered statistically significant in these analyses. We evaluated the importance of each selected feature in the Cox-PH model by their HR p-values. HR describes the relative contribution of a feature to the patient’s PI. HRs above one increase the risk, whereas below one decrease the risk. In the context of our model, HRs above one shorten the PE diagnosis time, while HRs below one lengthen it.

## Results

### Study design and dataset overviews

The overall study design is shown in **Figure 1**. The discovery cohort was extracted from patient records in the UM Health System from 2015 to 2022 with ICD-10 code access. All patients with a PE diagnosis after 20 weeks gestation were included in the cohort, and other exclusion criteria are detailed in Methods. The finalized UM discovery cohort consists of EMR records from 1178 pregnancies. Using the same inclusion and exclusion criteria, 881 pregnancies were identified in the validation dataset from UF. The patient characteristics for each cohort are listed in **Table 1**. The average maternal age of the discovery cohort was 30·2 and 29·1 in the validation cohort. The mean PE onset gestation age for this cohort was 251 days, and 257 for the validation cohort. We constructed and validated two models using this data: 1) a baseline model utilizing only patient medical history, demographics, and any new medical issues within the first 20 weeks of gestation; 2) a full model including those features from the baseline model, as well as additional information on medication, labs, and vitals within the first 20 weeks of pregnancy.

**Table 1:** Summaries of the characteristics of the study population. Data are presented as average (standard deviation) or counts (percentage % in the cohort).

| Characteristic | Discovery Cohort (N=1178) | Validation Cohort (N=881) |
| --- | --- | --- |
| **Maternal Age** | 30.2(5.67) | 29.1(6.18) |
| **Gravidity** | 2.31(1.74) | 2.82(2.04) |
| **Parity** | 0.68(1.12) | 1.17(1.5) |
| **Number of Fetuses** | 1.07(0.26) | 1.04(0.22) |
| **PE onset gestational age (days)** | 251(25.4) | 257(25.9) |
| **Current Smoker** | 61(5.2%) | 112(13%) |
| **Current Alcohol User** | 311(26%) | 184(21%) |
| **Race/Ethnicity** |  |  |
| African American | 195(17%) | 335(38%) |
| Asian | 74(6.3%) | 19(2.2%) |
| Hispanic | 58(4.9%) | 4(0.5%) |
| **History of PE** | 184(16%) | 117(13%) |
| **History of PE diagnosed in second trimester** | 66(5.6%) | 3(0.3%) |
| **Medical History** |  |  |
| Uncomplicated Type I Diabetes | 34(2.9%) | 19(2.2%) |
| Uncomplicated Type II Diabetes | 62(5.3%) | 22(2.5%) |
| Uncomplicated Hypertension | 201(17%) | 81(9.2%) |
| Kidney Disease | 14(1.2%) | 1(0.11%) |
| Clinical Diagnoses within 20 weeks gestation |  |  |
| Depression | 265(22%) | 19(2.2%) |
| Mood/Anxiety Disorder | 318(27%) | 0 |

**Figure 1.**
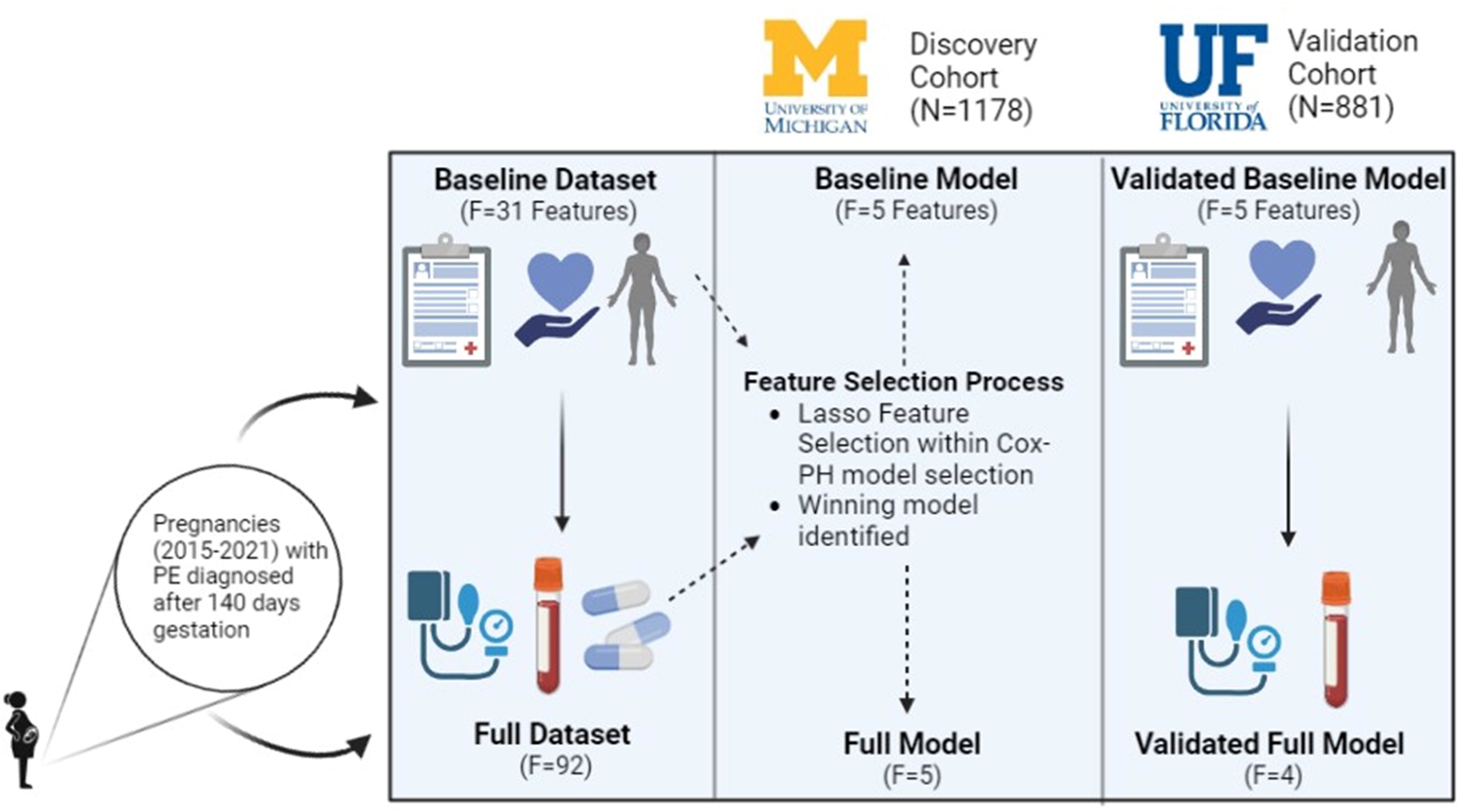
Study design and workflow. The discovery cohort was obtained from the University of Michigan Health System and a validation cohort of similar size and time was obtained from the University of Florida Health System. We constructed two PE predictive models: baseline and full model. The input variables in baseline models include patients’ demographics, lifestyle, comorbidities and medical history (N=31) and were reduced down to five features. The input for the full model includes additional lab tests and vital signs around PE diagnosis time, in addition to the variables in the baseline models (N=92) and was reduced down to five features for the discovery cohort, and four features for the validation cohort. We trained the Cox-PH models with LASSO regularization, using 80% training from the University of Michigan discovery cohort. We tested it on 20% hold-out data from the same discovery cohort, and validated it using the University of Florida validation cohort.

### Baseline model to predict PE onset time

We first built a baseline model using medical history, demographics, and medical information generated during the first 20 weeks of pregnancy. To build and test the model, we randomly split the data into an 80/20 ratio for training and testing datasets. We built the Cox-PH model with LASSO regulation with the UM training data under five-fold cross validation. We then applied this model to the 20% UM holdout testing data. The C-indices for the training and hold-out testing data of the baseline model are 0·62 and 0·64, respectively. On the external UF validation cohort, this model produced an C-index of 0·61, confirming its validity. Five features were selected for the baseline model following LASSO feature selection. Their respective hazard ratios (HR) and rankings in the multivariate Cox-PH are depicted in **Figure 2A and Table 2**. All features have positive HRs, showing that the presence of each feature increases PE risk and shortens the onset time of PE. These features arranged by their HRs, are: number of fetuses in pregnancy of interest (HR=25·17, p-value=1·84e-13), parity (HR=2·08, p-value=2·01e-06), history of uncomplicated hypertension (HR=2·01, p-value=1·01e-14), history of uncomplicated type II diabetes (HR=1·87, p-value=1·65e-05), and a mood/anxiety disorder (HR=1·24, p-value=0·00453).

**Table 2:**
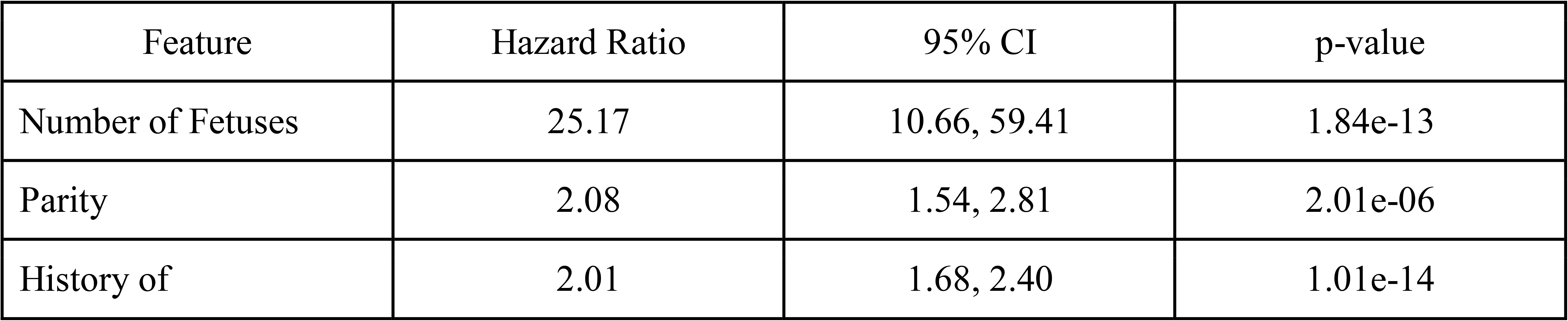
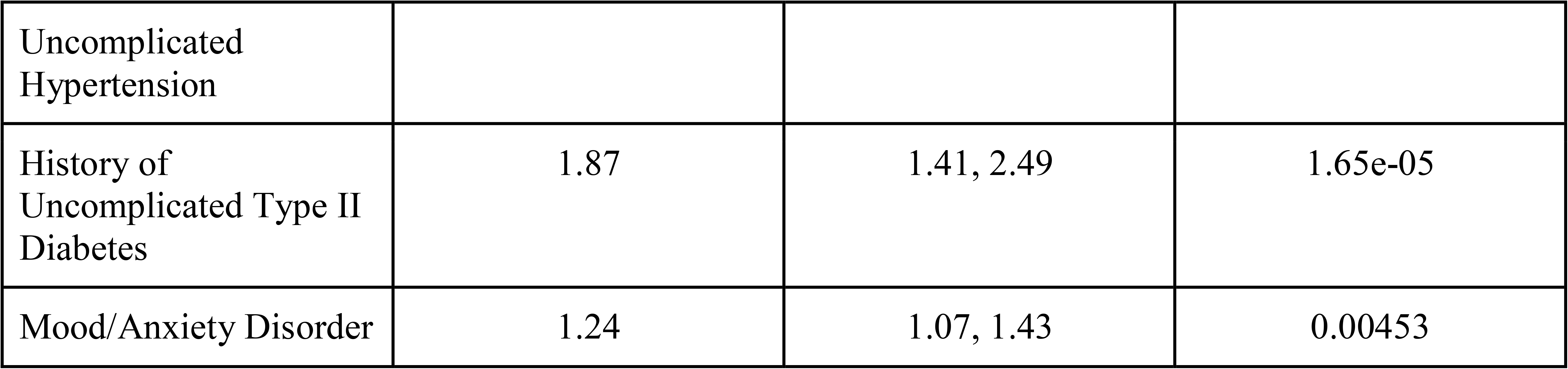
The selected features in the baseline model to predict PE onset time.

**Figure 2.**
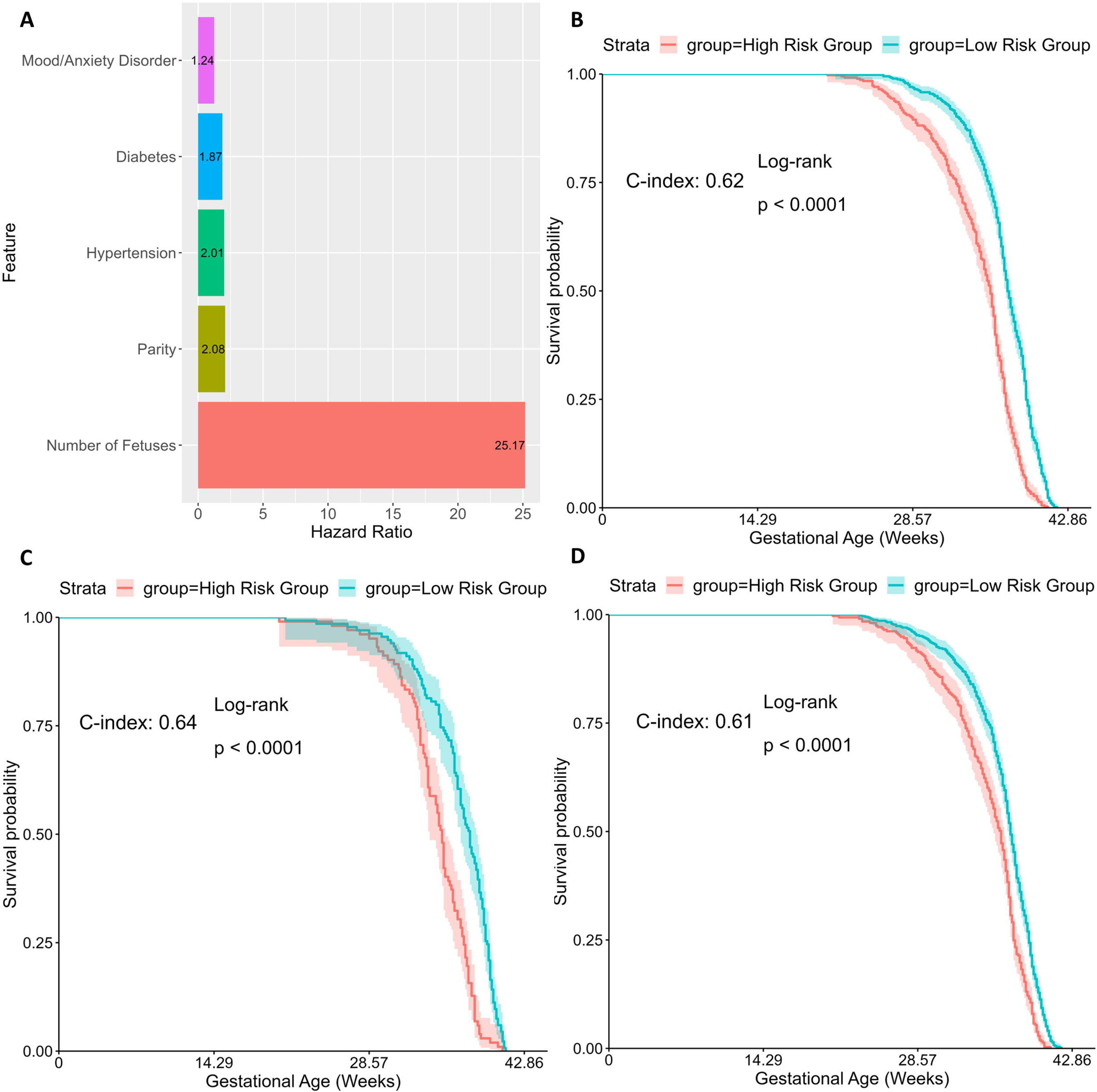
PE diagnosis time baseline model features and performance. (A) Bar plot of Hazard Ratios of the selected features by Cox-PH method with Lasso regularization. (B-D) Kaplan-Meier survival curves of high-risk (red) and low-risk (blue) pregnancies in the respective datasets. (B): UM training dataset; (C) hold-out testing set; (D) UF validation dataset.

In order to evaluate the discriminative power of this model, we dichotomized the patients from the training dataset into high and low risk groups by stratifying the samples using the median of the predicted prognosis index (PI=1·17) from the model. The two risk groups showed significant differences in prognosis (**Figure 2B, Supplementary Table 2**). The high-risk group was characterized by higher parity and number of fetuses, while the low-risk pregnancies had no prevalence of hypertension or diabetes (p-values: <2·26e-16 and 1.51e-13, respectively). We also applied the median PI value above to categorize samples into high vs. low risk groups in the hold-out (PI=1·17) and validation data (PI=2·38), similar to others.^17–20^ As shown in **Figure 2C** and **2D**, the Kaplan-Meier curves on these two risk groups are also significantly different (log-rank p-value <0·001).

### Full model for PE onset gestational age prediction

We next evaluated the addition of labs, vitals, and medication information (in the first 20 weeks) to the clinical data used in the baseline model. We constructed the new Cox-PH model the same way as done in the baseline model and obtained a five-feature Cox-PH model (**Figure 3A**). We name this feature-augmented model as the ‘full model’. It reaches C-indices of 0·66 and 0·69 for the training and hold-out testing datasets, respectively. This model also yields a C-index of 0·61 on the UF validation cohort, despite missing one of the five features (NSAID prescription) in the UF cohort. All five features have positive HRs (**Figure 3A** and **Table 3**). In descending order of HR, these features are: maximum diastolic blood pressure (HR=21·72, p-value=2·12e-09), number of fetuses in current pregnancy (HR=21·11, p-value=3·38e-15), parity (HR=1·81 p-value=3·08e-05), history of uncomplicated hypertension (HR=1·79, p-value=1·62e-12), and NSAID prescription (HR=1·35, p-value=0·00022). Three of these features, namely the number of fetuses, history of uncomplicated hypertension, and parity features are also selected previously by the baseline model (**Figure 3B**). Their HRs across the baseline and full models remain very similar, suggesting that they are all important in predicting PE onset time regardless of the other additional input information. Maximum diastolic blood pressure and NSAID prescription record are newly selected features unique to the full model (**Figure 3A, 3B**). The highest HR value from maximum diastolic blood pressure confirms its importance in prognostic prediction (**Figure 3A**).

**Table 3:** The selected features in the full model to predict PE onset time.

| Feature | Hazard Ratio | 95% CI | p-value |
| --- | --- | --- | --- |
| Max Diastolic Blood Pressure | 21.72 | 7.93, 59.74 | 2.12e-09 |
| Number of Fetuses | 21.11 | 9.88, 45.08 | 3.38e-15 |
| Parity | 1.81 | 1.37, 2.39 | 3.08e-05 |
| History of Uncomplicated Hypertension | 1.79 | 1.53, 2.11 | 1.62e-12 |
| NSAID Use | 1.35 | 1.15, 1.58 | 0.00022 |

**Figure 3.**
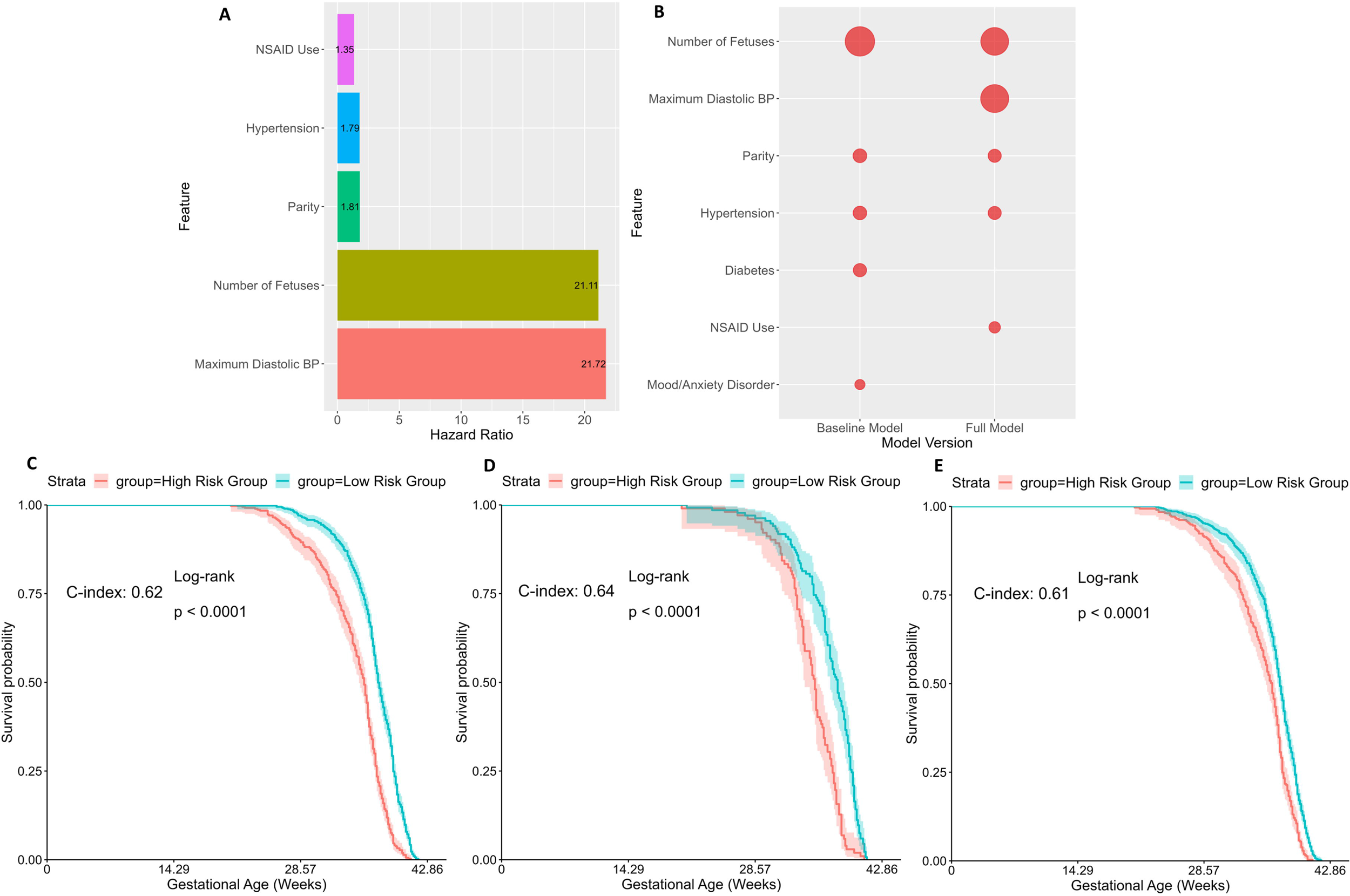
PE diagnosis time full model features and performance. (A) Bar plot of Hazard Ratios of the selected features in the full model by Cox-PH method with Lasso regularization. (B) The bubble plot of important features from PE baseline and full models. The size of bubbles represents the HR of each feature. (C-E) Kaplan-Meier survival curves of high-risk (red) and low-risk (blue) pregnancies in the respective datasets. (C): UM training dataset; (D) hold-out testing set; (E) UF validation dataset.

Similar to the baseline model, we stratified the patients into high vs. low risk groups using the median predicted PI value 5·15 from the training dataset (**Figure 3C**). The high-risk group was characterized by higher parity, higher number of fetuses and higher maximum diastolic blood pressure (**Supplementary Table 2)**. In contrast, the low risk group had no history of hypertension and rare use of NSAID medication. Blood pressure had the most statistically significant difference (p-value <2·2e-16), as expected. We applied the same median threshold to the 20% hold-out testing dataset (PI=5·08) and validation data (PI=5·18) for dichotomization (**Figures 3D, 3E**). Kaplan-Meier curves on these two risk groups in the testing set have even more significant differences in their diagnosis time (log-rank p-value <0·001).

## Discussion

This paper is the first of its kind to implement and validate a prognosis predicting model for PE onset time, rather than risks of PE, as many previous models do. It utilizes a comprehensive list of medical history, patient demographics, pregnancy characteristics, and medications, labs, and vitals collected in the first twenty weeks of pregnancy, which is the period before the symptoms of PE are manifested clinically. The models here not only confirm the importance of some previously known risk factors, such as number of fetuses, history of hypertension and parity, but also assign quantitative scores (weights) on the relative importance of these risk factors. More importantly, it identifies additional alarming factors to be considered in predicting PE onset time, such as the mood/anxiety disorder and the use of NSAID use during the first 20 weeks of pregnancy. The fact that maximum diastolic blood pressure had the highest hazard ratio in the full model confirms the importance of monitoring blood pressure as early as possible, even before PE is diagnosed clinically.

Besides the findings above, the models proposed here are relatively simple and straightforward, containing five important features for each model. This will reduce the burden of data collection and facilitates risk stratifications for clinicians in a clinical setting. The validation of these models’ performance on a second dataset from a vastly different institution - with different protocols, data collection, and data storage - demonstrates their potential to be generalized to different health institutions across the United States. The risk stratification of PE through this model provides clinicians with greater insight into the impact of these features on the development of PE.

Several areas of improvement may be considered in the future. Besides EMR, other information such as genetics, genomics, proteomics and metabolomics using maternal blood samples^21^ may help improve the prediction of onset time of PE. Additionally, lab tests that are typically administered later in pregnancy^22^, such as aspartate aminotransferase and alanine aminotransferase tests, are worthy of investigation for their prognostic values if they are measured within the first twenty weeks. Lastly, the current Cox-PH model is not designed to include longitudinal observations, limiting the kind of input variables to be incorporated in the model. Future work may benefit from more sophisticated modeling approaches^22^.

In conclusion, this study reports prognosis models to predict the onset gestational age of PE with EMR data prior to the first 20 weeks of pregnancy. They identify clinical and physiological factors for clinicians to watch out for that would increase a patient’s risk for earlier PE development.

## Data Availability

Data sharing is restricted. Access can be requested via the IRBs.

## Data Availability

Data sharing is restricted. Access can be requested via the IRBs.

## Declaration of Interests

The authors have no conflicts of interest to declare.

## List of Abbreviations

(PE): Preeclampsia
(EMRs): Electronic medical records
(C-index): Concordance index
(EOPE): Early-onset preeclampsia
(LOPE): Late-onset preeclampsia
(Cox-PH): Cox proportional hazards
(ICD): International Classification of Diseases
(BMI): Body mass index
(UF): University of Florida
(HR): Hazard Ratio
(PI): Prognosis Index
(WBCC): White Blood Cell Count
(MCHC): Mean corpuscular hemoglobin concentration
(MPV): Mean platelet volume
(NSAIDs): Nonsteroidal anti-inflammatory drugs

## Notes

### Competing Interest Statement

The authors have declared no competing interest.

### Funding Statement

LXG was supported by grants K01ES025434 awarded by NIEHS through funds provided by the trans-NIH Big Data to Knowledge (BD2K) initiative (www.bd2k.nih.gov), R01 LM012373 and LM012907 awarded by NLM, R01 HD084633 awarded by NICHD. DJL was supported by the National Institute of Diabetes and Digestive and Kidney Diseases (K01DK115632) and the University of Florida Clinical and Translational Science Institute (UL1TR001427). AM is supported by the National Center for Advancing Translational Science (5TL1TR001428). Additionally, the research reported in this publication was supported by the National Center for Advancing Translational Sciences of the National Institutes of Health under the University of Florida Clinical and Translational Science Award UL1TR001427.

### Author Declarations

The Institutional Review Board of the University of Michigan Medical School (HUM#00168171) gave ethical approval for this work. The Institutional Review Board of the University of Florida (#201601899) gave ethical approval for this work.

### Summary of Updates

Updated author order

## References

1. Bc Y, Rj L, Sa K. Pathogenesis of preeclampsia. Annual review of pathology. 2010;5. doi:10.1146/annurev-pathol-121808-102149

2. Al-Jameil N, Khan FA, Khan MF, Tabassum H. A Brief Overview of Preeclampsia. J Clin Med Res. 2013;6(1):1–7. doi:10.4021/jocmr.v6i1.1682

3. Lc C, s D, Pt S, et al. Diagnostic accuracy of placental growth factor in women with suspected preeclampsia: a prospective multicenter study. Circulation. 2013;128(19). doi:10.1161/CIRCULATIONAHA.113.003215

4. Wainstock T, Sergienko R, Sheiner E. Who Is at Risk for Preeclampsia? Risk Factors for Developing Initial Preeclampsia in a Subsequent Pregnancy. J Clin Med. 2020;9(4):1103. doi:10.3390/jcm9041103

5. Sibai BM, Stella CL. Diagnosis and management of atypical preeclampsia-eclampsia. Am J Obstet Gynecol. 2009;200(5):481.e1-7. doi:10.1016/j.ajog.2008.07.048

6. Haile DB, Aguade AE, Fetene MZ. Joint modeling of hypertension measurements and time-to-onset of preeclampsia among pregnant women attending antenatal care service at Arerti Primary Hospital, North Shoa, Ethiopia. Cogent Public Health. 2022;9(1):2022846. doi:10.1080/2331205X.2021.2022846

7. von Dadelszen P, Magee LA, Roberts JM. Subclassification of preeclampsia. Hypertens Pregnancy. 2003;22(2):143–148. doi:10.1081/PRG-120021060

8. Yang X, Ballard HK, Mahadevan AD, et al. Deep learning-based prognosis prediction among preeclamptic pregnancies using electronic health record data. Published online April 5, 2022:2022.04.03.22273366. doi:10.1101/2022.04.03.22273366

9. Elixhauser Comorbidity Software Refined for ICD-10-CM. Accessed February 7, 2023. https://www.hcup-us.ahrq.gov/toolssoftware/comorbidityicd10/comorbidity_icd10.jsp

10. Bernard N, Forest JC, Tarabulsy GM, Bujold E, Bouvier D, Giguère Y. Use of antidepressants and anxiolytics in early pregnancy and the risk of preeclampsia and gestational hypertension: a prospective study. BMC Pregnancy and Childbirth. 2019;19(1):146. doi:10.1186/s12884-019-2285-8

11. R: The R Project for Statistical Computing. Accessed February 7, 2023. https://www.r-project.org/

12. dplyr package - RDocumentation. Accessed February 7, 2023. https://www.rdocumentation.org/packages/dplyr/versions/1.0.10

13. gtsummary package - RDocumentation. Accessed February 7, 2023. https://www.rdocumentation.org/packages/gtsummary/versions/1.6.3

14. Friedman J, Hastie T, Tibshirani R, et al. glmnet: Lasso and Elastic-Net Regularized Generalized Linear Models. Published online November 27, 2022. Accessed February 7, 2023. https://CRAN.R-project.org/package=glmnet

15. mice package - RDocumentation. Accessed February 7, 2023. https://www.rdocumentation.org/packages/mice/versions/3.15.0

16. cindex function - RDocumentation. Accessed February 9, 2023. https://www.rdocumentation.org/packages/pec/versions/2022.05.04/topics/cindex

17. Ness RB, Roberts JM. Heterogeneous causes constituting the single syndrome of preeclampsia: a hypothesis and its implications. Am J Obstet Gynecol. 1996;175(5):1365–1370. doi:10.1016/s0002-9378(96)70056-x

18. English F, Kenny L, McCarthy F. Risk factors and effective management of preeclampsia. Integrated blood pressure control. 2015;8:7–12. doi:10.2147/IBPC.S50641

19. E P, S P, Tf M, D P, A N, Kh L. Clinical risk factors for preeclampsia in the 21st century. Obstetrics and gynecology. 2014;124(4). doi:10.1097/AOG.0000000000000451

20. Allahyari E, Foroushani A, Zeraati H, Mohammad K, Taghizadeh Z. A Predictive Model for the Diagnosis of Preeclampsia. JOURNAL OF REPRODUCTION AND INFERTILITY. Published online 2010. Accessed February 7, 2023. https://www.semanticscholar.org/paper/A-Predictive-Model-for-the-Diagnosis-of-Allahyari-Foroushani/e6f962dd332b1662b1f7c6455c52d58743acdfd4

21. Benny PA, Alakwaa FM, Schlueter RJ, Lassiter CB, Garmire LX. A review of omics approaches to study preeclampsia. Placenta. 2020;92:17-27. doi:10.1016/j.placenta.2020.01.008

22. Tarca A, Romero R, Benshalom-Tirosh N, et al. The prediction of early preeclampsia: Results from a longitudinal proteomics study. PLOS ONE. 2019;14:e0217273. doi:10.1371/journal.pone.0217273

