## Supplemental Tables for "Building and validating 5-feature models to predict preeclampsia onset time from electronic health record data" (from another version of this preprint: version 1, 2023-03-24)

**Supplementary Tables**

Supplementary Table 1: Inclusion/Exclusion Criteria

| Medical Condition | Inclusion/Exclusion | ICD Codes |
| --- | --- | --- |
| HELLP Syndrome | Exclusion | O14.2* |
| Pre-existing hypertension with superimposed preeclampsia | Exclusion | O11.* |
| Postpartum Preeclampsia | Exclusion | O14.95 |
| Preeclampsia | Inclusion | O14.0*, O14.1*, O14.9* |

Supplementary Table 2: Mean Feature Difference across Risk Groups for Baseline and Full Model Testing Datasets

| Model | Feature | Low-Risk | High-Risk | Welch Two-sample p-value |
| --- | --- | --- | --- | --- |
| Baseline Model | Number of Fetuses | 1.00 | 1.13 | <2e-16 |
|  | Parity | 1.14 | 2.11 | <2e-16 |
|  | Hypertension | 0.00 | 0.41 | <2e-16 |
|  | Uncomplicated Type II Diabetes | 0.00 | 0.13 | 1.51e-13 |
|  | Mood/Anxiety Disorder | 0.21 | 0.36 | 2.62e-07 |
| Full Model | Number of Fetuses | 1.00 | 1.10 | 5.57e-16 |
|  | Parity | 1.13 | 1.82 | <2e-16 |
|  | Hypertension | 0.00 | 0.34 | <2e-16 |
|  | Maximum Diastolic Blood Pressure | 71.31 | 82.82 | <2e-16 |
|  | NSAID Use | 0.01 | 0.39 | <2e-16 |
